# Parents’ intention to vaccinate their child for COVID-19: a cross-sectional survey (CoVAccS – wave 3)

**DOI:** 10.1101/2022.05.20.22275350

**Authors:** Louise E. Smith, Susan M. Sherman, Julius Sim, Richard Amlôt, Megan Cutts, Hannah Dasch, Nick Sevdalis, G James Rubin

## Abstract

**Objectives:** To investigate UK parents’ vaccination intention at a time when COVID-19 vaccination was available to some children.

**Study design:** Data reported are from the second wave of a prospective cohort study.

**Methods:** Online survey of 270 UK parents (conducted 4-15 October 2021). At this time, vaccination was available to 16- and 17-year-olds and had become available to 12- to 15- year-olds two weeks prior. We asked participants whose child had not yet been vaccinated how likely they were to vaccinate their child for COVID-19. Linear regression analyses were used to investigate factors associated with intention. Parents were also asked for their main reasons behind vaccination intention. Open-ended responses were analysed using content analysis.

**Results:** Parental vaccination intention was mixed (likely: 39.3%, 95% CI 32.8%, 45.7%; uncertain: 33.9%, 27.7%, 40.2%; unlikely: 26.8%, 20.9%, 32.6%). Intention was associated with: parental COVID-19 vaccination status; greater perceived necessity and social norms regarding COVID-19 vaccination; greater COVID-19 threat appraisal; and lower vaccine safety and novelty concerns. In those who intended to vaccinate their child, the main reasons for doing so were to protect the child and others. In those who did not intend to vaccinate their child, the main reason was safety concerns.

**Conclusions:** Parent COVID-19 vaccination and psychological factors explained a large percentage of the variance in vaccination intention for one’s child. How fluctuating infection rates, more children being vaccinated, and the UK’s reliance on vaccination as a strategy to live with COVID-19 may impact parents’ intention to vaccinate their child requires further study.

## Introduction

Vaccination has been one of the cornerstones of the public health response to COVID-19. However, there has been some debate over the need to vaccinate children due to the relatively lower severity of infection in this age group.^1^ While vaccination has been available in the UK for those aged 16 years and above since December 2020, vaccination has subsequently been extended to younger age groups (Box 1).

Box 1. Timeline of recommendations for COVID-19 vaccination in children and adolescents in England.

2 December 2020. MHRA gives approval for Pfizer/BioNTech vaccine in people aged 16 years and above.^2^

4 June 2021. MHRA gives approval for Pfizer/BioNTech vaccine in 12- to 15-year-olds.^3^

19 July 2021. JCVI advises that children and young people aged 12 years and above with specific underlying health conditions that put them at serious risk of COVID-19 be offered COVID-19 vaccination (2 doses, with an 8-week gap).^4^

4 August 2021. JCVI advises that all adolescents aged 16 to 17 years receive first dose of Pfizer/BioNTech vaccine.^5^

17 August 2021. MHRA gives approval for Moderna vaccine for 12- to 17-year-olds.^6^

2 September 2021. Half of 16- and 17-year-olds had received first COVID-19 vaccine.^7^

3 September 2021. JCVI release a report on vaccinating healthy 12- to 15-year-olds saying that “the health benefits from COVID-19 vaccination to healthy children aged 12 to 15 years are marginally greater than the potential harms … the margin of benefit is considered too small to support universal COVID-19 vaccination for this age group at this time”.^8^

13 September 2021. Children aged 12- to 15-years old in England will be offered one dose of Pfizer/BioNTech vaccine, following advice from the four UK Chief Medical Officers.^9^

14 September 2021. JCVI recommends a third dose (booster) for 16- to 49-year-olds with underlying health conditions.^10^

20 September 2021. NHS starts vaccine rollout in schools to children aged 12- to 15-year-olds.^11^

15 November 2021. JCVI recommends a second vaccine dose for all 16- to 17-year-olds (12 weeks or more after first dose).^12^

25 November 2021. European Medicines Agency recommends approval for Pfizer/BioNTech vaccine for 5- to 11-year-olds.^13^

29 November 2021. JCVI advises that all 12- to 15-year-olds should be offered a second dose of Pfizer-BioNTech vaccine.^14^

13 December 2021. NHS COVID-19 vaccine pass for international travel introduced for 12- to 15- year-olds.^15^

22 December 2021. MHRA gives approval for Pfizer vaccine for 5- to 11-year-olds.^16^

22 December 2021. JCVI recommends that children aged 5 to 11 years in a clinical risk group be offered COVID-19 vaccination (2 doses, with an 8-week gap). JCVI recommends that 16- to 17- year-olds and 12- to 15-year-olds in a clinical risk group or who are a household contact of someone who is immunosuppressed be offered a booster dose of the Pfizer-BioNTech vaccine no sooner than 3 months after they complete their primary vaccination course.^17^

16 February 2022. JCVI advises a “non-urgent offer” of vaccination to all children aged 5 to 11 years (two doses of Pfizer-BioNTech vaccine, at least 12 weeks apart).^18^

Abbreviations: JCVI = Joint Committee on Vaccination and Immunisation, MHRA = Medicines and Healthcare products Regulatory Agency, NHS = National Health Service, UK = United Kingdom, UKHSA = UK Health Security Agency.

Multiple factors are important in parents’ vaccine decision making for their children. Previous research indicates that uptake of child vaccination is associated with: perceived vaccine safety, generally positive vaccine attitudes, perceived susceptibility of the child to infection and greater social norms for vaccination.^19, 20^ Much research has been carried out on parents’ COVID-19 vaccine intention for their children, including two systematic reviews and meta-analyses. The first review searched papers published up to November 2021 (29 studies included; one conducted in the UK),^21^ while the second searched papers published up to December 2021 (44 studies included; two conducted in the UK; data collected March 2020 to September 2021).^22^ Both UK studies presented data collected before child vaccination was available in the UK.^23, 24^ Findings indicate that parental intention to vaccinate one’s child for COVID-19 is associated with psychological factors including greater perceived risk of COVID-19, more positive vaccine attitudes, and greater perceived safety of COVID-19 vaccination.^21, 22^ Novelty and a perceived lack of evidence about the effectiveness of child vaccination were associated with decreased parental intention to vaccinate one’s child.^21^ Evidence for associations with parent and child sociodemographic characteristics were mixed, but parental COVID-19 vaccine uptake (or intention) was consistently associated with intention to vaccinate one’s child.^21, 22^

This study aimed to investigate: UK parents’ intention to vaccinate their children against COVID-19 at a time when vaccination was available for some children; parents’ main reasons behind their intention; and factors associated with intention.

## Methods

This study reports data from the COVID-19 vaccination acceptability study (CoVAccS). Methods have been reported in more detail elsewhere.^25^

### Design

This formed part of a prospective cohort study (T1: 13-15 January 2021, T2: 4-15 October 2021). For this study, we report only data collected at T2.

### Participants

To be eligible for this study, participants had to be aged 18 years or over, live in the UK and have not completed our previous study (CoVAccS 1), due to similarities in survey materials.^26^ 1500 participants completed the first wave of data collection (T1), of whom 1148 also completed the second wave (response rate 76.5%; T2). For this study, we included participants if they indicated that they were the parent or legal guardian of a dependent child aged 17 years or younger (n=270).

### Measures

Full survey materials are available online.^27^ Unless otherwise specified, all items were measured at T2.

Parents were asked to complete the survey with one of their children in mind (index child). We asked parents to think about their child who had most recently had a birthday.

#### Personal and clinical characteristics

At T1, all participants were asked to provide information about their sex, age, ethnicity, religion, highest qualification, employment status, total household income, and region where they live. At T2, we asked whether they had a chronic illness and recoded people as being “at risk” of COVID-19 based on NHS guidance.^28^

At T2, parents were also asked for characteristics of the index child, namely their sex, age, whether they were their first child, whether the child had previously had COVID-19, and whether they had a chronic illness (recoded to “at risk” of COVID-19 based on NHS guidance^28^).

#### Psychological factors

Items were informed by psychological theories and psychosocial factors known to affect vaccine uptake.^26, 29^ Parents were asked a series of nine statements about their beliefs about COVID-19 and vaccination with reference to their child on an 11-point scale from “strongly disagree (0)” to “strongly agree (10)”. Items asked about perceived risk of COVID-19 to the child, worry about the child catching COVID-19, perceived susceptibility to, and severity of,

COVID-19 for the child, anticipated regret about the child catching COVID-19 if they had not been vaccinated, belief that the vaccine could give the child COVID-19, regretting vaccination if the child were to experience side effects, worry about the safety of COVID-19 vaccination for children, and perceived social norms about child vaccination.

Parents were also asked about their agreement that vaccination is generally a good thing.

#### Vaccination uptake

Parents were asked if their child had been vaccinated against COVID-19. Response options were: “yes, they’ve had one dose”, “yes, they’ve had two doses”, “no”, “don’t know”, and “prefer not to say”.

Parents who indicated that they had not vaccinated their child at the time of data collection were asked how likely it was that their child would have a COVID-19 vaccine, eventually. Statements differed based on the age of the child, due to the differing availability of the vaccine by child age (likelihood “if a coronavirus vaccination becomes available to your child” [parents of children aged 0 to 11 years], likelihood “when a coronavirus vaccination becomes available to your child” [parents of children aged 12 to 15 years], likelihood “now that a coronavirus vaccination is available to your child” [parents of children aged 16 to 17 years]).

Parents whose children had not yet been vaccinated were also asked to report the main reason why their child was “likely or unlikely to have a coronavirus vaccination”.

### Ethics

We obtained ethical approval for this study from Keele University’s Research Ethics Committee (reference: PS-200129).

### Power

We conducted a post-hoc power analysis based on linear regression analyses. With an achieved sample size of 220, a 1% two-tailed significance level, and testing 16 predictors, we had 88.8% power to detect medium effect sizes (*f*^2^=0.15).

### Analysis

Parents who reported that their child had been vaccinated were excluded from analyses of vaccination intention.

We categorised parents as being very likely to vaccinate their child if they scored 8 to 10 (on the 0-10 point numerical response scale), very unlikely to vaccinate their child if they scored 0 to 2, and uncertain about vaccinating their child if they scored 3 to 7 (*a priori* cut-offs based on our previous work^26, 29^). To investigate whether parents’ vaccine intentions differed by index child age (0 to 11 years, 12 to 15 years, 16 to 17 years), we used a one-way ANOVA.

We conducted a principal components analysis to aid feature identification for psychological items. As components were thought likely to be correlated, we used oblique (direct oblimin) rotation. All psychological factor items pertaining to parents’ beliefs about COVID-19 illness and vaccination in their child were included in the analysis.

Factors associated with parents’ intention to vaccinate one’s child were investigated using a linear regression analysis (n=224), using the full eleven-point response scale as the outcome measure. Variables were entered into the regression model in blocks; the order was selected *a priori* based on theoretical relevance (block 1: parent socio-demographic characteristics, block 2: child socio-demographic characteristics, block 3: general vaccination beliefs and attitudes, block 4: beliefs and attitudes about COVID-19 and vaccination). To control the rate of Type 1 errors in the regression analysis, we set statistical significance at *p*≤.01 and therefore calculated 99% confidence intervals (CIs) for regression coefficients.

Reasons behind parents’ intention to vaccinate their child for COVID-19 were analysed qualitatively using content analysis in which emerging codes were identified from the data.^30^

## Results

### Participant characteristics

The 270 participants included in the study were mostly female (54.8%, n=148/270), white (85.6%, n=231/270), with a mean age of 42.1 years (SD=9.3; Table 1).

**Table 1.**
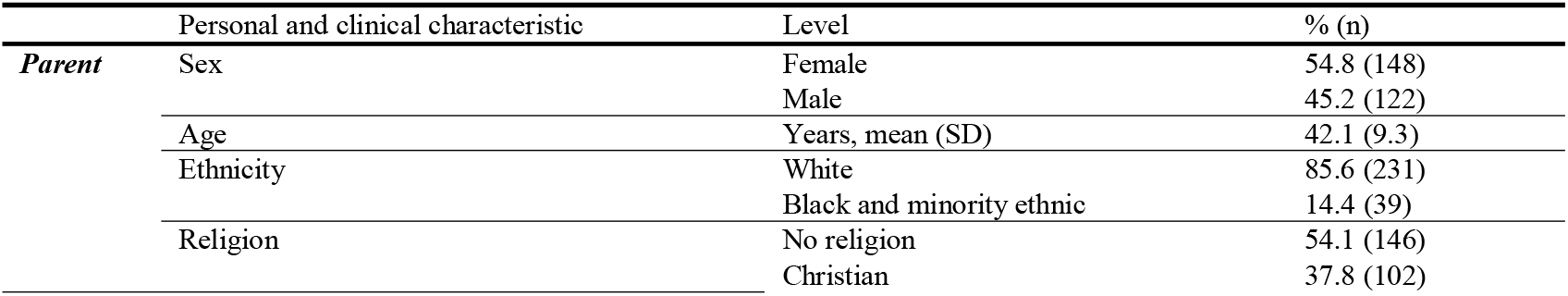
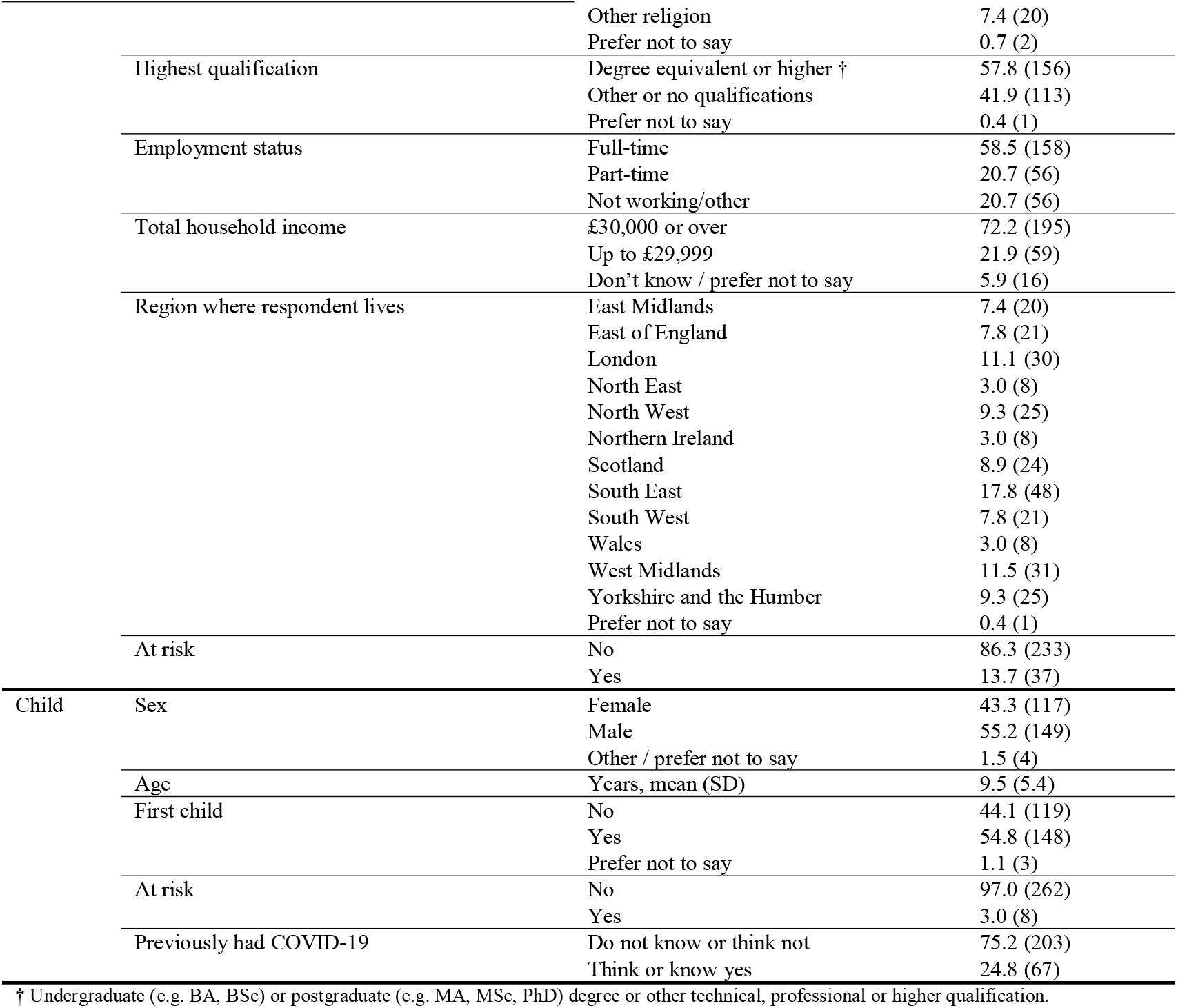
Participant characteristics. Data are frequencies (percentages), except for age: mean (SD). Data are for all parents regardless of the vaccination status of their child (total *n*=270).

### Vaccine uptake

Parent-reported child vaccine uptake was low, with 3.0% (n=8, 95% CI 1.5% to 5.7%) indicating that their child had had two doses of the vaccine, 14.1% (n=38; 95% CI 10.4% to 18.7%) reporting that their child had had one dose of the vaccine, and 83.0% (n=224, 95% CI 78.0% to 87.0%) reporting that their child had not received the vaccine.

### Vaccine intention

Parents who had not yet vaccinated their child were split in their intention to vaccinate their child (Figure 1). Of parents of children of all ages, 39.3% (n=88/224, 95% CI 33.1% to 45.8%) were likely to vaccinate their child, 33.9% (n=76/224, 95% CI 28.0% to 40.4%) were uncertain, and 26.8% (n=60/224, 95% CI 21.4% to 32.9%) were unlikely to vaccinate their child. The mean (SD) parental intention score on the 0–10 scale was 5.62 (3.68), and was similar in relation to children in different age bands: 5.38 (3.59) for those aged 0–11; 6.29 (3.77) for those aged 12–15; 5.40 (4.07) for those aged 16–17.

**Figure 1.**
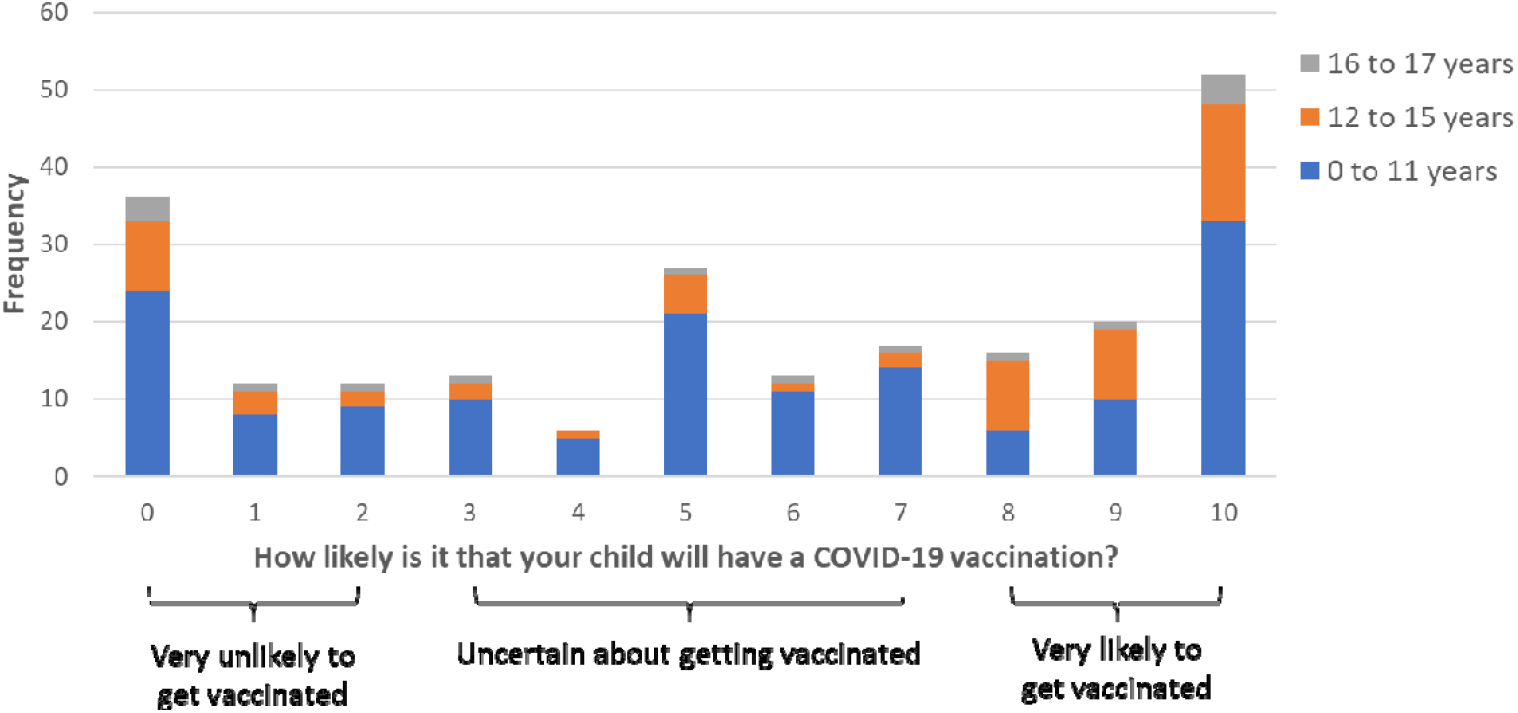
Perceived likelihood of child having a vaccination (0=“extremely unlikely” to 10=“extremely likely”) by child age, with cut-off points used to categorise participants’ vaccination intention.

### Principal components analysis

Three components emerged from the principal components analysis on beliefs and attitudes about COVID-19 and vaccination, accounting for 72% of the variance in the original items (see supplementary materials). One component related to illness beliefs and attitudes (‘COVID-19 threat appraisal’), and two components related to vaccination beliefs and attitudes (‘perceived necessity and social norms of child COVID-19 vaccination’ and ‘vaccine safety and novelty concerns’).

### Factors associated with parental vaccination intention

219 participants had complete data and were included in regression analyses. The overall regression model explained 66.9% of the variance, with parents’ socio-demographic characteristics and beliefs and attitudes about COVID-19 and vaccination explaining the largest percentage of variance. Vaccination intention was associated with: having had a COVID-19 vaccine oneself; greater perceived necessity and perceived social norms regarding COVID-19 vaccination; greater COVID-19 threat appraisal; and lower vaccine safety and novelty concerns (Table 2).

**Table 2.** Results of the full linear regression model analysing associations with parental vaccination intention (adjusted *R*^2^ = 0.669). Parameter estimates relate to the full model containing all predictors. The unstandardized regression coefficients represent the change in parental vaccination intention for a one-unit increase in the predictor variable (or, for dummy variables, a shift from the reference category to the category concerned). The model was based on 219 study participants with complete data.

| Predictor variable | Level | Standardized coefficient | Unstandardized coefficient | 99% confidence interval | *p* value |
| --- | --- | --- | --- | --- | --- |
| **Block 1 – parent personal and clinical characteristics <sup>a</sup>** |  |  |  |  |  |
| Sex (reference: female) | Male | -0.036 | -0.264 | -1.171, 0.643 | 0.450 |
| Age (years) |  | 0.026 | 0.011 | -0.050, 0.072 | 0.639 |
| Ethnicity (reference: white) | Black and minority ethnic | 0.097 | 1.061 | -0.237, 2.359 | 0.035 |
| Religion (reference: none) | Christian | 0.003 | 0.025 | -0.809, 0.858 | 0.938 |
|  | Other | -0.034 | -0.538 | -2.434, 1.357 | 0.461 |
| Qualifications (reference: other) | Degree equivalent or higher | 0.006 | 0.044 | -0.812, 0.900 | 0.894 |
| Employment status (reference: not working/other) | Part-time | 0.076 | 0.693 | -0.508, 1.894 | 0.135 |
|  | Full-time | 0.026 | 0.195 | -0.863, 1.254 | 0.632 |
| At risk – self (reference: no) | Yes | -0.104 | -1.158 | -2.340, 0.025 | 0.012 |
| Vaccinated for COVID-19 – self (reference: no) | Yes | 0.157 | 1.695 | 0.336, 3.054 | 0.001* |
| **Block 2 – child personal and clinical characteristics <sup>b</sup>** |  |  |  |  |  |
| Sex (reference: female) | Male | -0.013 | -0.096 | -0.895, 0.703 | 0.755 |
| Age (years) |  | -0.043 | -0.032 | -0.138, 0.073 | 0.428 |
| First child (reference: not) | Yes | 0.040 | 0.303 | -0.552, 1.158 | 0.357 |
| Had covid before (reference: think not) | Yes | 0.067 | 0.600 | -0.350, 1.550 | 0.102 |
| At risk – child (reference: no) | Yes | -0.001 | -0.033 | -3.092, 3.025 | 0.977 |
| **Block 3 – general vaccination beliefs and attitudes <sup>c</sup>** |  |  |  |  |  |
| In general, vaccination is a good thing (0–10) | 0 = strongly disagree, 10 = strongly agree | 0.125 | 0.246 | -0.009, 0.500 | 0.013 |
| **Block 4 – beliefs and attitudes about COVID-19 and vaccination <sup>d</sup>** |  |  |  |  |  |
| Component: perceived necessity and social norms of child COVID-19 vaccination |  | 0.506 | 1.884 | 1.396, 2.372 | <0.001* |
| Component: COVID-19 threat appraisal |  | 0.141 | 0.521 | 0.089, 0.953 | 0.002* |
| Component: vaccine safety and novelty concerns |  | -0.277 | -1.020 | -1.430, -0.610 | <0.001* |

\* $p \leq .01$ ; <sup>a</sup> variables in this block explained 29.2% of the variance; <sup>b</sup> variables in this block explained a further 0.6% of the variance; <sup>c</sup> variables in this block explained a further 7.1% of the variance; <sup>d</sup> variables in this block explained a further 30.0% of the variance.

### Reasons behind intention

The main reasons behind vaccination intention in parents who were likely to vaccinate their child were to protect the child, to protect others, and because the child had chosen to (Table 3). The main reasons behind intention in parents who were unlikely to vaccinate their child were safety concerns, feeling that the threat to the child of COVID-19 was low, and that there was no personal need for the child to be vaccinated.

**Table 3.**
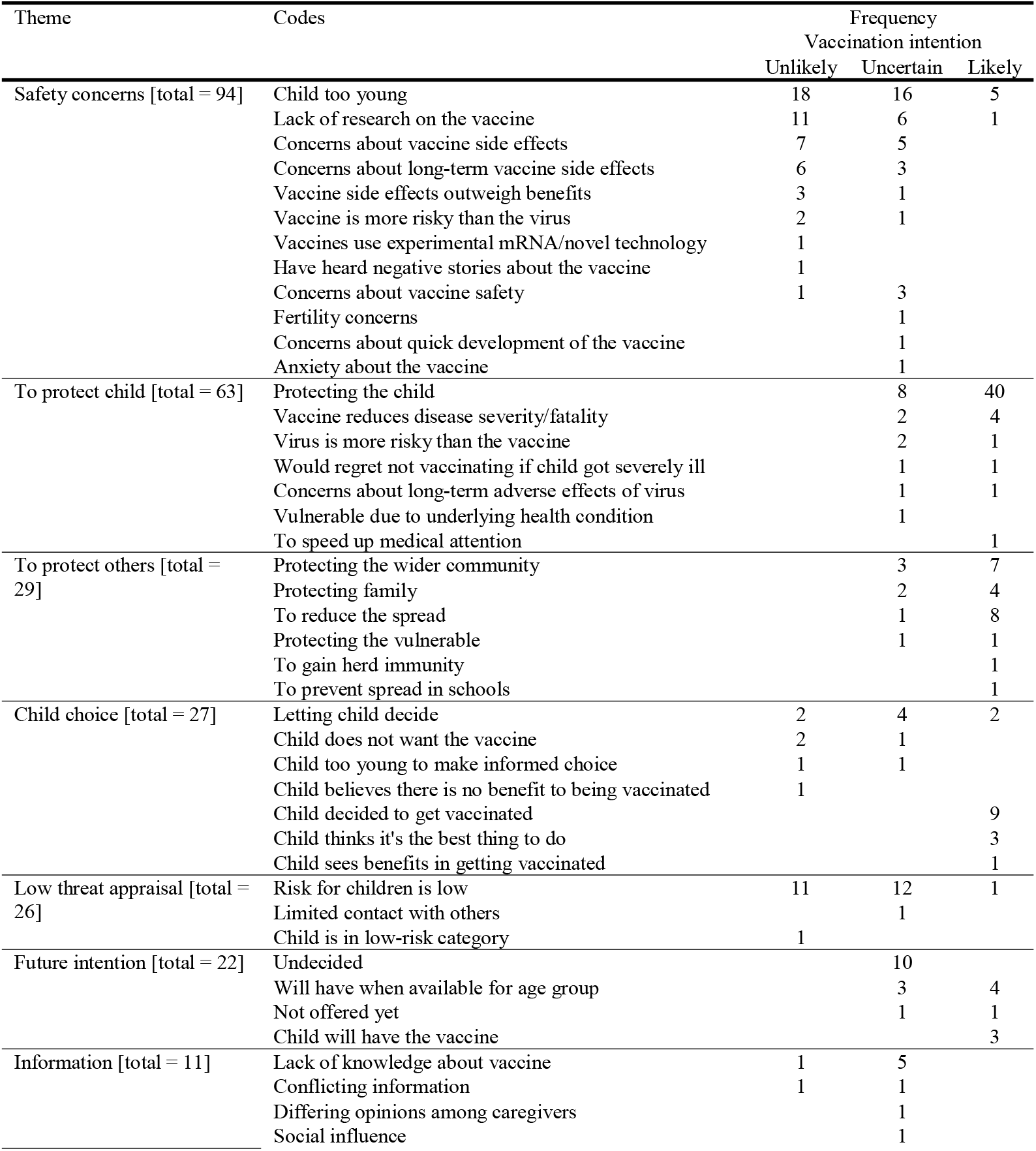
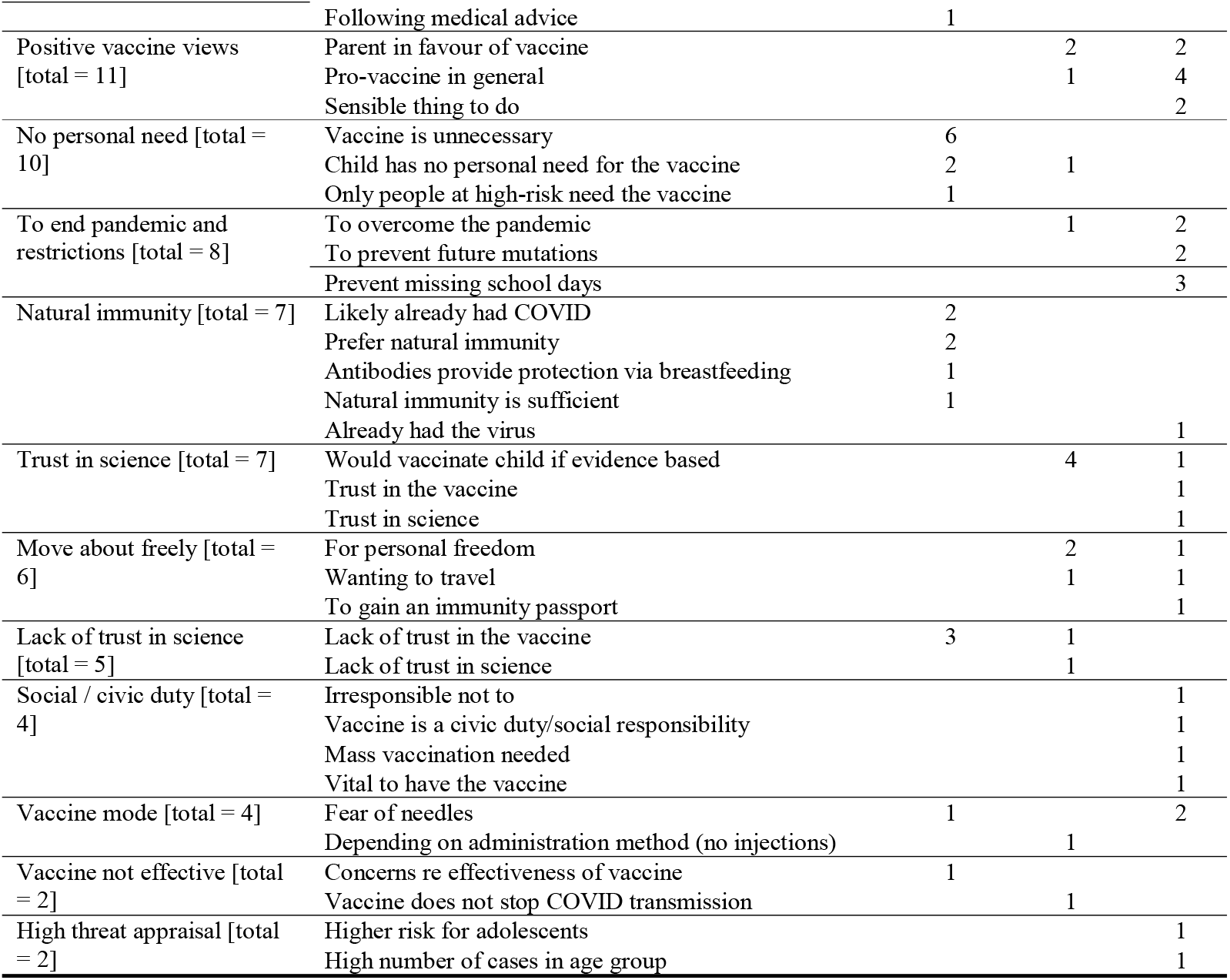
Thematic categorization of codes generated by content analysis of reasons for or against vaccinating one’s child, by vaccination intention. Themes are presented in descending order of overall frequency.

## Discussion

By the end of data collection (15 October 2021), COVID-19 vaccination was available to all 16- and 17-year-olds and had recently become available to 12- to 15-year olds in England (20 September 2021^11^; Box 1). Official NHS figures indicate that by this time, 1,487,232 cumulative doses of COVID-19 vaccine had been given to under 18s (data up to 15 October 2021, n=1,234,290 first dose, n=251,825 second dose, n=1,117 booster doses^31^). This equates to approximately 9% of the English population under 18 (14,191,190 aged 17 years and under^32^). Uptake in our sample of parents was slightly higher, with approximately 17% indicating that their child had received at least one dose.

Of those who had not yet vaccinated their child, parents’ vaccination intentions were mixed, with 39% indicating that they were likely to vaccinate their child, 34% being unsure, and 27% being unlikely. The percentage of those likely to vaccinate their child is lower than recent meta-analytic results, which found a willingness rate of approximately 60%.^21, 22^ It is also lower than other UK parental vaccination studies. One study of English parents with a child aged 18 months or younger found that 48% definitely would accept a COVID-19 vaccine for their child (a further 41% “unsure but leaning towards yes”; data collected 19 April to 11 May 2020).^23^ UK data from a multi-country survey found that 64% of women who were either pregnant at the time of data collection or who had one child aged 18 years or under were likely to vaccinate their child (data collected 28 October to 18 November 2020).^24^ The discrepancy between these findings and our results may be because of the increased debate surrounding child COVID-19 vaccination,^1^ and decreased perceived risk of COVID- 19 between January and October 2021,^25^ itself associated with vaccination intention.^20^

Having had a COVID-19 vaccine oneself was strongly associated with parents’ vaccine intentions for their child, in line with systematic review results.^21, 22^ This is the same pattern of results seen in previous pandemics, with intention to be vaccinated for pandemic influenza being associated with previous seasonal influenza vaccination during the 2009/2010 H1N1 influenza pandemic.^33^ Parental socio-demographic characteristics explained 29% of the variance in child vaccination intention in our study, with parental vaccination being the only individual variable associated, suggesting that this variable exerts a strong influence on parental vaccination intention.

We found no other associations between parental vaccination intention for their child and parent or child sociodemographic characteristics. This is likely due to low power to detect smaller effects. Official UK figures show there are differences in uptake of vaccination in children aged 12 to 15 years by sociodemographic variables.^34^ Namely, vaccine uptake is higher in children who are of Chinese and Indian ethnicities, live in less deprived areas, speak English as a first language, do not receive free school meals, and do not have special educational needs.

Beliefs and attitudes about COVID-19 and vaccination explained a further 30% of the variance in parents’ vaccination intentions. We found that parental vaccination intention was associated with greater perceived necessity of COVID-19 vaccination and social norms (believing that most other children will receive a COVID-19 vaccine), greater perceived risk of COVID-19, and greater perceived safety of vaccination. This is in line with other results found during the COVID-19 pandemic,^21, 22^ routine childhood vaccination,^19, 20^ and theories of uptake of health behaviours (e.g. the Protection Motivation Theory^35^). High case numbers in primary and secondary school age children, such as those seen in September to November 2021 and January 2022,^36^ may also affect parents’ vaccination intention, through perceived susceptibility to infection. Factors associated with parental vaccination intention were similar to those associated with uptake of COVID-19 vaccination by oneself.^25^

Parents who intended to vaccinate their child most commonly cited the protection of the child and others as the main reasons behind their intentions. A similar pattern of results has been found in children aged 12 to 18 years (school years 7 to 13 in the UK), with the most common reasons for deciding to have a COVID-19 vaccine being to protect oneself and other people from getting COVID-19, and because the rest of the family had been vaccinated.^37^ For those who did not intend to vaccinate their child, the main reasons were safety concerns and not perceiving COVID-19 to be a great threat to their child. This reflects findings from the Office for National Statistics, in which 24% parents of children aged 5 to 11 years were unlikely to agree to their having a COVID-19 vaccine, with the main reasons behind this lack of vaccine intention being worry about side effects and waiting to see how the vaccine works.^37^

As the UK’s response to the pandemic shifts to “living with COVID-19,” this strategy is in part relying on adult vaccination as a means to reduce serious infection in the absence of other non-pharmaceutical interventions (testing, self-isolation, wearing a face covering, limits on social mixing).^38^ The risks and benefits of vaccination to children have been more balanced,^17^ leading to greater debate about whether children should be vaccinated.^1^ Parents need to be able to make an informed decision as to whether they vaccinate their child. Since data collection, COVID-19 vaccination has been approved and recommended in younger age groups (now offered to everyone aged 5-years and over; see Box 1). Factors affecting parents’ decision to vaccinate their child are numerous and likely interlinked.^19, 20^ While social norms for vaccination may increase with time as more children are vaccinated and perceptions of vaccine novelty may decrease, the landscape of the COVID-19 pandemic is constantly changing, with the risk of new variants and removal of restrictions. How this may affect parents’ perceived risk of their child being infected and, in turn, their vaccination intention and uptake remains to be understood.

We measured self-reported intention to vaccinate one’s child when the vaccine was not yet available to children of most age groups. While the study was well powered to detect medium effect sizes, we had limited power to detect smaller effect sizes. However, our regression model explained 67% of the variance in parents’ vaccination intention. This indicates good explanatory power for a study using these methods, with social science research and public opinion surveys typically giving low *R*^*2*^ values.^39, 40^ This study was part of a prospective cohort study. Participants recruited into the study at T1 were broadly representative of UK adults based on age, sex and ethnicity.^29^ Questions about child vaccination were only asked at T2, to those who completed the follow-up survey (response rate 76.5%) and who indicated that they were the parent or guardian of a child aged 17 years or under.^25^ We cannot be sure that the sample included in this survey is representative of UK parents.

### Conclusion

Parents’ COVID-19 vaccination intention for their child was mixed at a time when the vaccination was available for some children. Vaccination intention was associated with having been vaccinated for COVID-19 oneself, greater perceived necessity of, and social norms for, vaccination, greater perceived threat of COVID-19, and greater perceived safety of COVID-19 vaccination for children. Parents most commonly reported that they intended to vaccinate their child to protect the child and others, while the main reason behind not intending to vaccinate one’s child was due to safety concerns.

## Data Availability

Data are available online at

https://osf.io/tehg8/

## Sources of funding

Data collection was funded by a Keele University Faculty of Natural Sciences Research Development award to SMS, JS and NS, and a Kings COVID Appeal Fund award granted jointly to LS, GJR, RA, NS, SMS and JS. NS research is supported by the National Institute for Health Research (NIHR) Applied Research Collaboration (ARC) South London at King’s College Hospital NHS Foundation Trust. NS is a member of King’s Improvement Science, which offers co-funding to the NIHR ARC South London and is funded by King’s Health Partners (Guys and St Thomas NHS Foundation Trust, King’s College Hospital NHS Foundation Trust, Kings College London and South London and Maudsley NHS Foundation Trust), and the Guy’s and St Thomas’ Foundation. The views expressed are those of the authors and not necessarily those of the NIHR, the charities, UK Health Security Agency or the Department of Health and Social Care.

## Conflicts of Interest

NS is the director of the London Safety and Training Solutions Ltd, which offers training in patient safety, implementation solutions and human factors to healthcare organizations and the pharmaceutical industry. At the time of writing GJR is acting as an expert witness in an unrelated case involving Bayer PLC, supported by LS. LS, RA and GJR were members of the Scientific Advisory Group for Emergencies or its subgroups. The other authors have no conflicts of interest to declare.

## Author contributions

LS, JS, RA, NS, GJR and SMS conceptualized and acquired funding for the study. SMS programmed the survey, curated the data and was responsible for the administration of the project. LS, JS, MC and HD undertook formal analyses. LS wrote the original draft of the manuscript. JS, MC, HD, RA, NS, GJR and SMS reviewed and edited drafts.

## Data sharing

Data are available online.^27^

## Supplementary materials

### Results of principal components analysis

A scree plot identified that psychological items loaded onto three main components. The item “The coronavirus vaccination could give my child coronavirus” did not load on to any component. We therefore re-ran the principal components analysis excluding this variable. Table S1 shows item loadings on to components.

**Table S1.**
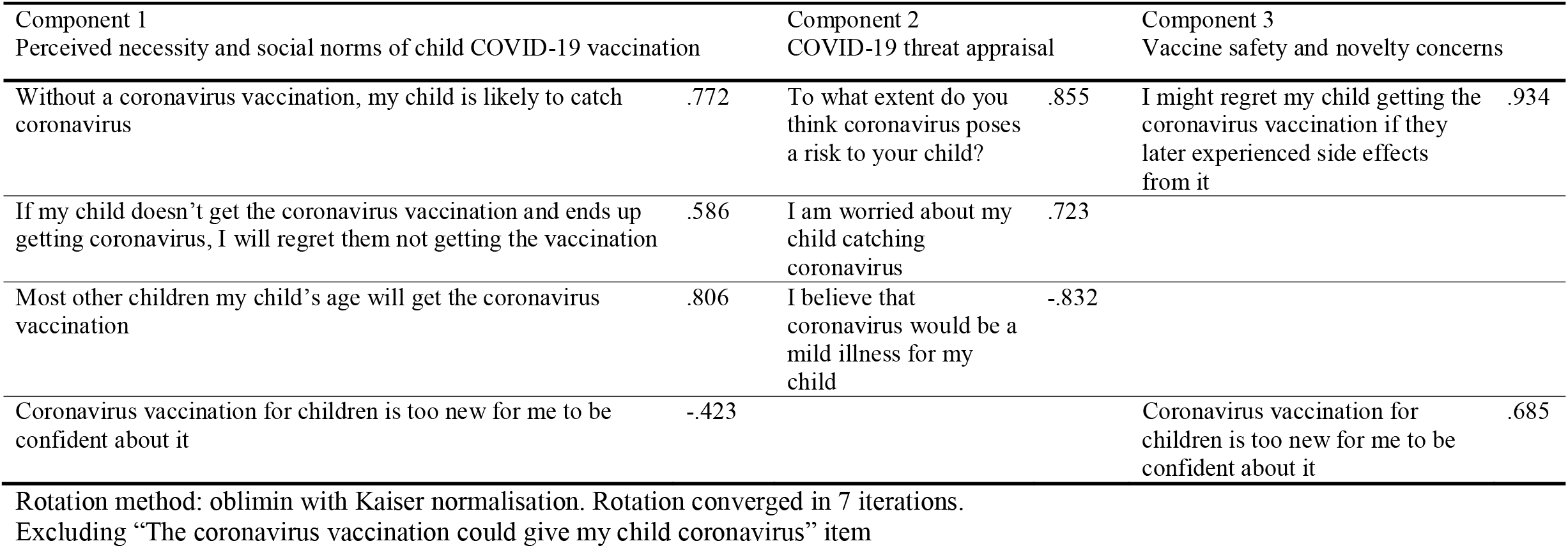
Loadings of items measuring psychological factors onto components identified (only loadings over .400 are shown).

